# Evaluation of VIDAS® diagnostic assay prototypes detecting dengue virus NS1 antigen and anti-dengue virus IgM and IgG antibodies

**DOI:** 10.1101/2021.02.19.21252059

**Authors:** Somphavanh Somlor, Ludovic Brossault, Marc Grandadam

## Abstract

Dengue is a serious tropical disease caused by the mosquito-borne dengue virus (DENV). Performant, rapid and easy-to-use assays are needed for the accurate diagnosis of acute DENV infection. We evaluated the performance of three prototype assays developed for the VIDAS^®^ automated platform to detect dengue NS1 antigen and anti-dengue IgM and IgG antibodies. Positive and negative agreement with competitor enzyme-linked immunosorbent assays (ELISA) and rapid diagnostic tests (RDT) was evaluated in 91 Lao patients (57 adults, 34 children) with acute DENV infection. The VIDAS^®^ NS1 assay showed the best overall agreement (95.6%) with the competitor NS1 ELISA. Both VIDAS^®^ NS1 and NS1 ELISA assays also demonstrated high sensitivity relative to DENV RNA RT-PCR set as gold standard (85.7% and 83.9%, respectively). In contrast, NS1 RDT was less sensitive relative to DENV RNA RT-PCR (72.7%). The overall agreement of VIDAS^®^ IgM and IgG assays with the competitor assays was moderate (72.5% for IgM ELISA, 76.9% for IgG ELISA, and 68.7% for IgM and IgG RDT). In most analyses, test agreements of the VIDAS^®^ assays were comparable in adults and children. Altogether, the VIDAS^®^ dengue prototypes performed very well and appear to be suitable for routine detection of dengue NS1 antigen and anti-dengue IgM/IgG antibodies.

## 1. Introduction

Dengue is a mosquito-borne viral disease caused by infection with any of four dengue virus (DENV) serotypes (DENV-1 to DENV-4) [1]. With an estimated 390 million annual infections in 2010, of which 96 million with clinical manifestations, dengue is a major public health burden, notably in tropical and sub-tropical regions of the world [2,3]. Its incidence dramatically increased in the last 50 years in these regions and has recently begun to increase in Europe [1,3,4]. Dengue is a systemic and progressive disease with symptoms ranging from mild to severe [1,3]. Following an early febrile phase that lasts for 2 to 7 days, patients enter a critical phase during which patients’ condition either improves or worsens, and may evolve into life-threatening severe dengue, with a potentially fatal outcome [1,3]. DENV serotype and secondary infection are among the factors associated with disease severity [1]. If the patient survives the first 24-48-hour critical phase, the final recovery phase will take its course over the following 48-72 hours [1,3]. No specific therapy against dengue exist but patient management protocols have been implemented to limit severe dengue complications [1,3]. The first licensed dengue vaccine is restricted to people aged 9-45 years and is only recommended for those with evidence of a past dengue infection [5].

In this context, rapid and performant approaches are urgently needed to better prevent and control dengue outbreaks, notably via improved epidemiologic surveillance, and to better manage dengue-infected patients, via early diagnosis. Early dengue diagnosis based on clinical manifestations is difficult as the symptoms of the febrile phase do not differentiate from those of unrelated febrile diseases and do not distinguish between mild and severe dengue progression [1,3]. Several diagnostic methods have been developed, combining virological and serological assays. Their application depends on the time after onset of symptoms. During the early (acute) phase of infection, circulating DENV RNA can be detected by reverse-transcription PCR (RT-PCR) while the secreted viral non-structural protein 1 (NS1) antigen can be detected by immunoassays such as enzyme-linked immunosorbent assay (ELISA) or immuno-chromatographic rapid diagnostic tests (thereafter referred to as RDT). In the post-acute phase, the detection of anti-DENV immunoglobulin M (IgM) and immunoglobulin G (IgG) produced in response to infection, using ELISA- or RDT-based serological assays can support the diagnosis of a DENV infection. The current guidelines (WHO, CDC, PAHO) recommend a RT-PCR and/or NS1 antigen assay within the first 5 to 7 days of illness to confirm DENV infection [3,6,7]. Past this time, the detection of anti-DENV IgM and/or IgG is highly suggestive of a DENV infection [3,6,7]. The production of anti-DENV IgM being usually transient and that of IgG being sustained post infection, testing both anti-DENV IgM and IgG can improve the sensitivity of diagnosis [1,8–10]. Moreover, given the persistence of anti-DENV IgG after an infection, as opposed to anti-DENV IgM, which levels can vary greatly [11–13], the detection of high level IgG in the early acute phase supports a possible secondary DENV infection [1,8–10,14].

Altogether, it is now acknowledged that the combined testing of DENV RNA and/or NS1 antigen with anti-DENV IgM and IgG serology increases the sensitivity of DENV infection diagnosis, by broadening the time window of diagnosis after onset of fever [8,10,15–19]. Because RT-PCR assays are complex and costly, albeit being highly accurate, healthcare providers are turning to easy-to-use ELISA- and RDT-based assays detecting DENV NS1 antigen and anti-DENV IgM and IgG antibodies. ELISA and RDT present both pros and cons. In comparative studies, ELISA are usually more sensitive than RDT [8,12,19–22]. Anti-DENV IgM and IgG ELISA can show poor specificity due to cross-reactivity with antibodies directed against other flaviviruses [3,12,19,22,23]. RDT are rapid, and easier to perform and to implement in field settings. However, the visual interpretation of RDT results is operator-dependent and can be a drawback [24].

Altogether, an ideal DENV diagnostic assay should combine the performance accuracy of ELISA with the possibility to evaluate the best DENV diagnostic markers (NS1 antigen, and anti-DENV IgM and IgG antibodies), on a rapid, easy-to-use, and possibly automated readout system. The VIDAS^®^ dengue prototype assays (VIDAS^®^ DENGUE NS1 Ag, VIDAS^®^ Anti-DENGUE IgM and VIDAS^®^ Anti-DENGUE IgG) were developed to reach this goal. VIDAS^®^ is an automated platform now established worldwide for the accurate and rapid performance of antigen and serological diagnostic assays. This study aimed to evaluate the three VIDAS^®^ dengue prototype assays in 91 patients with acute DENV infection from Laos, a country highly endemic for all four DENV serotypes. We analysed the concordance of results of the three VIDAS^®^ assays to that of competitor ELISA assays (Dengue NS1 Antigen DxSelect™, Focus Diagnostics; Panbio Dengue IgM Capture ELISA and Panbio Dengue IgG Indirect ELISA, Abbott) and of a RDT (SD BIOLINE Dengue Duo, Dengue NS1 Ag + IgG/IgM, Abbott).

The VIDAS^®^ NS1 assay demonstrated high sensitivity relative to DENV RNA RT-PCR set as gold standard (85.7% [95% CI: 74.3-92.6%]) and strong overall agreement with the competitor NS1 ELISA (95.6% [95% CI: 89.1-98.8%]). In comparison, the overall agreement of the VIDAS^®^ NS1 assay with the NS1 RDT was lower (88.8% [95% CI: 80.5-93.8%]). The VIDAS^®^ IgM and IgG assays showed moderate overall agreement with the competitor assays (72.5% [95% CI: 62.6-80.6%] for IgM ELISA; 76.9% [95% CI: 67.3-84.4%] for IgG ELISA; 68.7% [95% CI: 58.1-77.6%] for IgM and IgG RDT). In most analyses, test agreements of the VIDAS^®^ assays were comparable in adults and children. Altogether, the VIDAS^®^ dengue prototypes appear to be suitable for routine detection of dengue NS1 antigen and anti-dengue IgM/IgG antibodies in both adults and children, in regions endemic for DENV.

## 2. Materials and Methods

### 2.1. Patients and samples

Plasma samples were collected between 2012 and 2019 as part of the dengue surveillance system established by the Institut Pasteur du Laos (Vientiane, Laos), where dengue is endemic. A total of 250 frozen EDTA plasmas from patients with suspected DENV infection were selected [25]. All plasmas were leftovers of samples initially used for routine laboratory tests scheduled for the diagnosis of suspected dengue cases, and approved for use for research purposes, including the development of diagnostic tools. This study was conducted according to the guidelines of the Declaration of Helsinki, and approved by the Institut Pasteur du Laos’ institutional review board and the National Ethic Committee for Health Research of the Ministry of Health of Lao PDR (No. 049 NECHR). Written informed consent was obtained from all adult patients and from a parent or legal guardian in the case of child patients. Patients were informed on the possible use of remaining samples and data, after anonymization, for research purposes.

Sera negative for dengue infection were collected from 51 healthy volunteers recruited by the French blood bank (Etablissement Français du Sang, EFS). Each healthy donor signed a written informed consent for the use of blood for research purposes. EFS obtained from the French ministry of research the authorization to collect and to transfer samples to partners (Ministère de l’Enseignement Supérieur, de la Recherche et de l’Innovation, reference AC-2017-2958).

### 2.2. Study design and definitions

This study aimed to evaluate and compare the diagnostic performance of VIDAS^®^ dengue prototype assays detecting DENV NS1 antigen (VIDAS^®^ DENGUE NS1 Ag) and anti-DENV IgM and IgG antibodies (VIDAS^®^ Anti-DENGUE IgM and VIDAS^®^ Anti-DENGUE IgG, respectively), to that of competitor assays. To be able to compare NS1, IgM and IgG assay performance (i.e. detecting both early and late markers) on the same set of samples, patients with acute DENV infection were included in the analysis (Figure 1). Dengue being hyper-endemic in Laos, the studied patient population was expected to include both primary and secondary DENV infections and encompass all four DENV serotypes. An acute DENV infection was defined as samples positive for DENV RNA and/or NS1 antigen. The presence of DENV RNA in plasmas was assessed by real-time RT-PCR, using a “universal” assay detecting all four DENV serotypes [26]. When possible, the serotype (DENV-1 to DENV-4) of the samples positive by universal RT-PCR was determined using specific primer pairs and probes described by Ito et al. [27] and adapted for a multiplex RT-PCR assay. The NS1 antigen assay applied to determine acute infection was the Dengue NS1 Antigen DxSelect™ (Focus Diagnostics; thereafter referred to as NS1 FOCUS), used as reference test.

**FIGURE 1.**
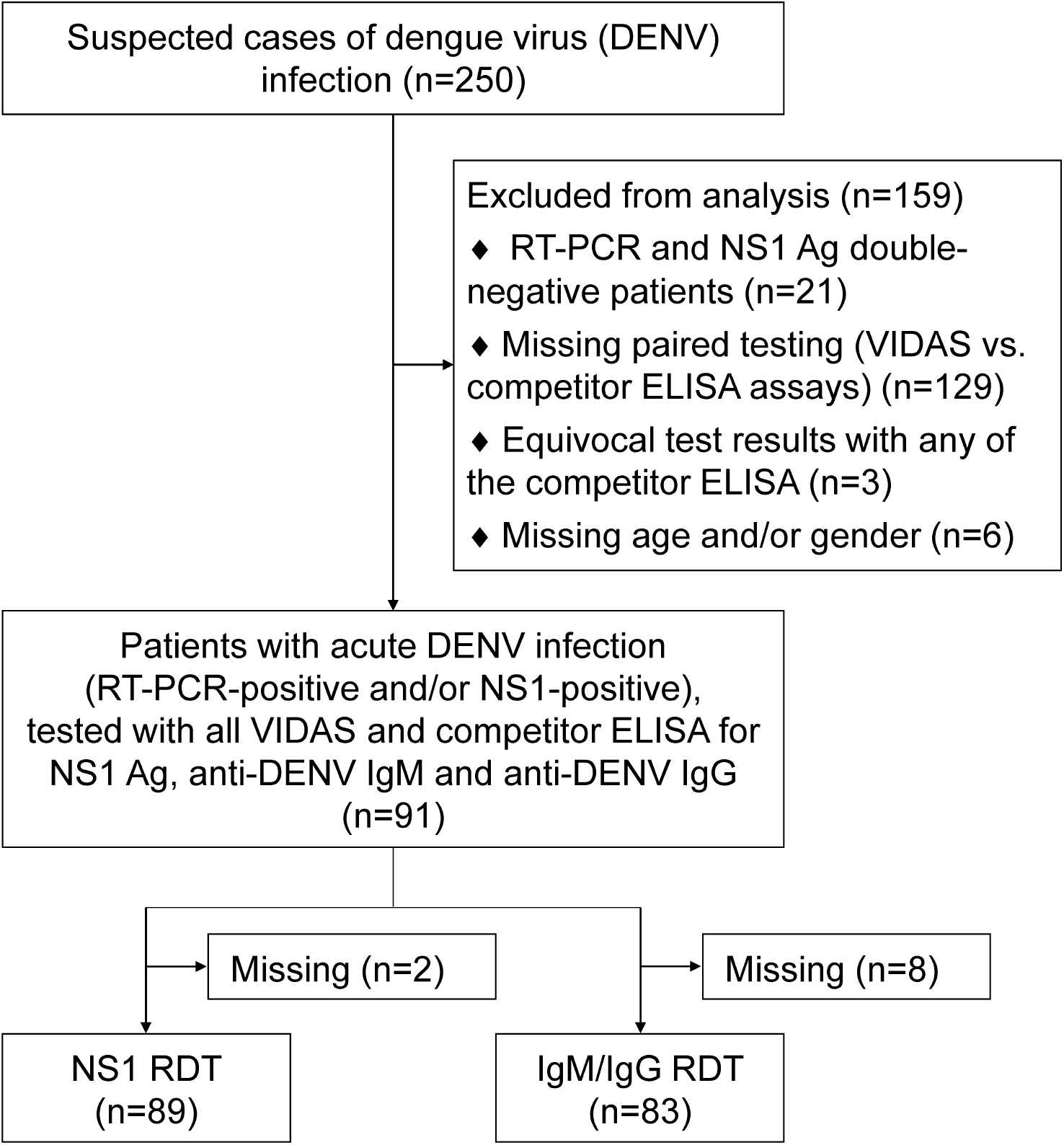
Study flow diagram. Out of 250 suspected cases of DENV infection, 91 identified as acute DENV infection (RT-PCR-positive and/or NS1 antigen-positive) were tested with the three VIDAS^®^ prototypes and competitor assays (ELISA and RDT). Of these 91 samples, 89 and 83 were also tested with a RDT specific for NS1 and IgM/IgG, respectively. Abbreviations: Ag, antigen; DENV, dengue virus; ELISA, enzyme-linked immunosorbent assay; IgM, immunoglobulin M; IgG, immunoglobulin G; NS1, non-structural protein 1; RDT, rapid diagnostic test; RT-PCR, real-time reverse-transcription polymerase chain reaction.

Selected samples were tested on the three VIDAS^®^ dengue prototype assays and with the Panbio Dengue IgM Capture ELISA and Panbio Dengue IgG Indirect ELISA (Abbott), in addition to the Dengue NS1 Antigen DxSelect™ (Focus Diagnostics) mentioned above. When possible, samples were also tested with the rapid diagnostic test (RDT) SD BIOLINE Dengue Duo, Dengue NS1 Ag + IgG/IgM (Abbott), thereafter referred to as NS1 RDT, IgM RDT and IgG RDT (Figure 1).

In addition, the three VIDAS^®^ dengue prototype assays were tested on sera from healthy volunteers (EFS blood donations) negative for all three competitor ELISA: Dengue NS1 Antigen DxSelect™ (Focus Diagnostics), Panbio Dengue IgM Capture ELISA and Panbio Dengue IgG Indirect ELISA (Abbott).

### 2.3. VIDAS^®^ assays

VIDAS^®^ dengue prototypes (VIDAS^®^ DENGUE NS1 Ag, VIDAS^®^ Anti-DENGUE IgM, VIDAS^®^ Anti-DENGUE IgG; bioMérieux, France) are automated qualitative two-step immunoassays combined with enzyme-linked fluorescent assay (ELFA) detection, developed for the VIDAS^®^ family of instruments. The Solid Phase Receptacle (SPR^**®**^) serves as the solid phase as well as the pipetting device. Reagents for the assay are ready-to-use and pre-dispensed in the sealed reagent strip. All steps are performed automatically by the instrument, within 40 to 60 minutes.

For the VIDAS^®^ DENGUE NS1 Ag assay, the Dengue NS1 antigen is captured by a monoclonal antibody specific for the four serotypes coated on the interior of the SPR wall. During the second step, the captured NS1 antigen is detected by monoclonal antibodies specific for the NS1 antigen of the four DENV serotypes conjugated to alkaline phosphatase.

For the VIDAS^®^ Anti-DENGUE IgM assay, total IgM are captured by a monoclonal antibody specific for human IgM coated on the interior of the SPR. During the second step, anti-DENV IgM are specifically detected by a recombinant tetravalent DENV envelope domain III (EDIIIT2) protein conjugated to alkaline phosphatase. The chimeric EDIIIT2 protein is composed of the antigenic, DENV-specific envelope domain III of the four DENV serotypes [28].

For the VIDAS^®^ Anti-DENGUE IgG assay, anti-DENV IgG are captured by the EDIIIT2 antigen coated on the interior of the SPR. During the second step, the captured anti-DENV IgG are detected by an antibody specific for human IgG conjugated to alkaline phosphatase.

During the final detection step of the three VIDAS^®^ dengue prototype assays, the substrate (4-Methyl-umbelliferyl phosphate) is cycled in and out of the SPR. The conjugate enzyme catalyzes the hydrolysis of the substrate into a fluorescent product (4-Methyl-umbelliferone). Fluorescence is measured at 450 nm and a relative fluorescence value (RFV) is generated (background reading subtracted from the final fluorescence reading). The results are automatically calculated by the instrument, according to a standard (S1), and an index value (i) is obtained (where i=RFV_sample_/RFV_S1_). The test is interpreted as negative when i < 1.0 and positive when i ≥ 1.0.

The positivity cut-off values for the Dengue NS1, IgM and IgG tests were determined based on area under the receiver operating characteristic (ROC) curve and Youden index analyses, using clinically characterized positive and negative human samples.

### 2.4. Competitor assays

The competitor ELISA used were: Dengue NS1 Antigen DxSelect™ (EL1510; Focus Diagnostics), Panbio Dengue IgM Capture ELISA (01PE20; Abbott) and Panbio Dengue IgG Indirect ELISA (01PE30; Abbott). Assays were conducted according to the manufacturer’s recommendations, except for the use of plasmas instead of sera. The use of plasma was not considered as a fundamental issue for this research study since the test results were not used for patient diagnosis. Of note, a study demonstrated comparable qualitative diagnostic interpretation of the Panbio Dengue IgM Capture ELISA and of the Panbio Dengue IgG Indirect ELISA whether using serum or plasma samples [29]. For the three competitor ELISA, a test was interpreted as negative when index values < 0.9 and positive when index values > 1.1. Samples with equivocal index values (between 0.9 and 1.1) were excluded from the analysis (Figure 1).

The RDT SD BIOLINE Dengue Duo, Dengue NS1 Ag + IgG/IgM (11FK46; Abbott) was performed and interpreted according to the manufacturer’s recommendations.

### 2.5. Statistical analysis

Statistical analyses were conducted using SAS version 9.4. Positive, negative and overall agreement were calculated between a reference test (competitor ELISA or RDT) and VIDAS^®^ Dengue tests, and the respective 95% confidence intervals (CI) were computed. Positive agreement between RT-PCR and the three Dengue NS1 assays was also evaluated (defined as sensitivity relative to RT-PCR set as gold standard). The McNemar test was used to compare the sensitivity of the three DENV NS1 assays. Differences in assay agreement between adults and children were evaluated using a chi-square test. Two-sided Fisher’s exact test was used alternatively in the case of too small sample sizes.

VIDAS^®^ Dengue index values were displayed as Tukey box plots using GraphPad Prism 5.04. Two-group comparisons of index value distributions between adults and children were performed using the nonparametric two-tailed Mann-Whitney *U*-test (MWU-test) with normal approximation. *P*-values ≤ 0.05 were considered statistically significant.

## 3. Results

### 3.1. Patients’ characteristics

Out of 250 plasma samples of patients with suspected DENV infection, 91 samples of acute DENV infection (RT-PCR-positive and/or NS1 FOCUS-positive) were tested with all three VIDAS^®^ Dengue prototype assays and all three competitor ELISA assays for dengue NS1 antigen, anti-DENV IgM and anti-DENV IgG antibodies. Of these 91 samples, 89 and 83 were also tested with an RDT specific for NS1 antigen and anti-DENV IgM/IgG, respectively (Figure 1).

Patients’ characteristics are shown in Table 1. Of the 91 acute dengue patients, 57 (62.6%) were adults, 51 (56.0%) were female, 56 (61.5%) were RT-PCR-positive and 82 (90.1%) were positive in the NS1 FOCUS ELISA. The median time to symptom onset (fever) was in the expected range given the acute phase of the disease (4.5 days in adults and 3.0 days in children). The number of serotype-specific RT-PCR determination was too low to perform a subgroup analysis (Table 1).

**Table 1.** Patients’ characteristics.

|  |  |
| --- | --- |
| Study population, N (%) | 91 (100.0%) |
| Adults | 57 (62.6%) |
| Children | 34 (37.4%) |
| Age in years, median (range) |  |
| Total | 21.0 (5-63) |
| Adults | 27.0 (19-63) |
| Children | 11.5 (5-18) |
| Gender, N (%) |  |
| Female | 51 (56.0%) |
| Male | 40 (44.0%) |
| Serotype, N (%) |  |
| DENV-1 | 3 (3.3%) |
| DENV-2 | 4 (4.4%) |
| DENV-3 | 10 (11.0%) |
| DENV-4 | 3 (3.3%) |
| Unknown | 71 (78.0%) |
| Time to symptom onset in days, median (range) |  |
| Total (N=87) | 4.0 (0.0-12.0) |
| Adults (N=54) | 4.5 (0.0-12.0) |
| Children (N=33) | 3.0 (0.0-8.0) |
| Unknown (N=4) | N/A |
| Acute infection status, N (%) |  |
| PCR+/NS1+ (FOCUS) | 47 (51.6%) |
| PCR-/NS1+ (FOCUS) | 35 (38.5%) |
| PCR+/NS1- (FOCUS) | 9 (9.9%) |

### 3.2. VIDAS^®^ index distribution

The distribution of index values generated by the three VIDAS^®^ prototype assays was comparable in adults and children (Figure 2; MWU-test, *P* > 0.05). The median (interquartile range) index values in the total population were 112.3 (63.9-142.1) for VIDAS^®^ DENGUE NS1 Ag (Figure 2A), 1.6 (0.7-4.8) for VIDAS^®^ Anti-DENGUE IgM (Figure 2B) and 2.2 (0.4-26.7) for VIDAS^®^ Anti-DENGUE IgG (Figure 2C).

**FIGURE 2.**
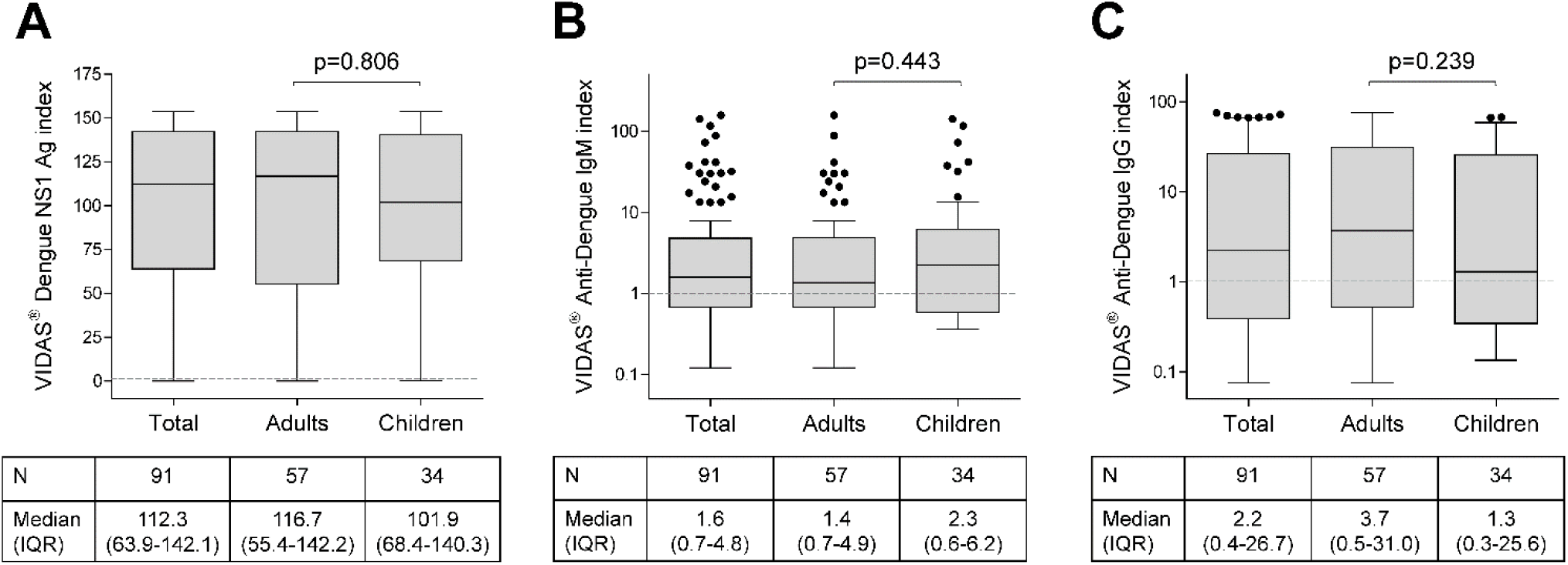
Distribution of index values generated by the VIDAS^®^ prototypes VIDAS^®^ DENGUE NS1 Ag (A), VIDAS^®^ Anti-DENGUE IgM (B) and VIDAS^®^ Anti-DENGUE IgG (C). Index values are depicted as Tukey box plots in the total population of patients with acute DENV infection (N=91), and as per adults (N=57) and children (N=34). The grey dashed line shows the positivity cut-off (i = 1.0). VIDAS^®^ Anti-DENGUE IgM and IgG index values (**B, C**) are shown in log10 scale to better visualize the index distribution. Index median and interquartile range (IQR) is shown below each graph. Differences in index distribution between adults and children were tested with the nonparametric two-tailed Mann-Whitney *U*-test; the differences were not statistically significant (*P* > 0.05 for the three VIDAS^®^ prototypes).

In line with these quantitative data, most of the comparative analyses of qualitative VIDAS^®^ results described below showed no significant differences between adults and children, as indicated in the respective tables.

### 3.3. Positive agreement of dengue NS1 antigen assays with RT-PCR as gold standard

Virus isolation is considered the “gold standard” of DENV infection diagnosis [7,30]. However, detection of DENV RNA by real-time RT-PCR, which is highly sensitive and specific [1,3,7,8,10], is often used in practice as a gold standard for acute DENV infection [21,31,32]. To evaluate the sensitivity of the different NS1 antigen assays, the positive agreement of the VIDAS^®^ DENGUE NS1 Ag, the NS1 FOCUS ELISA and the NS1 RDT with RT-PCR set as gold standard was evaluated in the 56 RT-PCR-positive patients (Table 2). The VIDAS^®^ DENGUE NS1 Ag assay and the NS1 FOCUS ELISA showed the highest sensitivity, with a positive agreement (95% confidence interval [CI]) with RT-PCR of 85.7% (74.3-92.6%) and 83.9% (72.2-91.3%), respectively (McNemar VIDAS^®^ vs. FOCUS, p=0.564). The NS1 RDT exhibited a lower sensitivity, with a positive agreement (95% CI) with RT-PCR of 72.7% (59.8-82.7%) (McNemar VIDAS^®^ vs. RDT, p=0.008).

**Table 2.** Positive agreement of the respective NS1 antigen assays with RT-PCR (N=56)

| Reference test | Population | VIDAS |  | FOCUS |  | RDT<sup>a</sup> |  |
| --- | --- | --- | --- | --- | --- | --- | --- |
|  |  | n/N | %<br>[95% CI] | n/N | %<br>[95% CI] | n/N | %<br>[95% CI] |
| RT-PCR | Total | 48/56 | 85.7%<br>[74.3-92.6] | 47/56 | 83.9%<br>[72.2-91.3] | 40/55 | 72.7%<br>[59.8-82.7] |
|  | Adults | 31/39 | 79.5%<br>[64.5-89.2] | 31/39 | 79.5%<br>[64.5-89.2] | 24/38 | 63.2%<br>[47.3-76.6] |
|  | Children | 17/17 | 100.0%<br>[80.5-100.0] | 16/17 | 94.1%<br>[73.0-99.0] | 16/17 | 94.1%<br>[73.0-99.0] |

<sup>a</sup>SD BIOLINE Dengue Duo Rapid Test (Dengue NS1 Ag); Fisher's Exact Test between adults and children: VIDAS® (p=0.090), FOCUS (p=0.250) and RDT (p=0.022); Abbreviations: CI, confidence interval; RDT, rapid diagnostic test.

### 3.4. Comparison of VIDAS^®^ DENGUE NS1 Ag assay with competitor assays

The positive agreement of the VIDAS^®^ DENGUE NS1 Ag to the NS1 FOCUS ELISA (used as reference test) was very high (97.6%; 95% CI [91.5-99.7]) (Table 3). This is in line with the comparably high sensitivity of both assays (Table 2). The negative agreement of the VIDAS^®^ NS1 assay with the NS1 FOCUS ELISA was 77.8% and the 95% CI was wide (45.3-93.7%). This result is difficult to interpret due to the small number of cases in this group (n=9). The overall agreement was very high with 95.6% (95% CI [89.1-98.8]).

**Table 3.** Concordance of VIDAS® Dengue NS1 Ag and competitor assays.

| Reference test | Population | Positive Agreement |  | Negative Agreement |  | Overall Agreement |  |
| --- | --- | --- | --- | --- | --- | --- | --- |
|  |  | n/N | %<br>[95% CI] | n/N | %<br>[95% CI] | n/N | %<br>[95% CI] |
| **Dengue NS1 Antigen DxSelect™ (Focus)** | Total | 80/82 | 97.6%<br>[91.5-99.7] | 7/9 | 77.8%<br>[45.3-93.7] | 87/91 | 95.6%<br>[89.1-98.8] |
|  | Adults | 48/49 | 98.0%<br>[89.1-99.9] | 7/8 | 87.5%<br>[52.9-97.8] | 55/57 | 96.5%<br>[87.9-99.6] |
|  | Children | 32/33 | 97.0%<br>[84.2-99.9] | 0/1 | 0.0%<br>[0.0-97.5] | 32/34 | 94.1%<br>[80.9-98.4] |
| **NS1 RDT<sup>a</sup>** | Total | 70/70 | 100.0%<br>[94.9-100.0] | 9/19 | 47.4%<br>[27.3-68.3] | 79/89 | 88.8%<br>[80.5-93.8] |
|  | Adults | 41/41 | 100.0%<br>[91.4-100.0] | 8/15 | 53.3%<br>[30.1-75.2] | 49/56 | 87.5%<br>[76.4-93.8] |
|  | Children | 29/29 | 100.0%<br>[88.1-100.0] | 1/4 | 25.0%<br>[4.6-69.9] | 30/33 | 90.9%<br>[76.4-96.9] |

<sup>a</sup>SD BIOLINE Dengue Duo Rapid Test (Dengue NS1 Ag); Fisher's Exact Test between adults and children on overall agreement: VIDAS® vs. FOCUS (p=0.628) and VIDAS® vs. RDT (p=0.739); Abbreviations: CI, confidence interval; RDT, rapid diagnostic test.

The positive agreement of the VIDAS^®^ DENGUE NS1 Ag to the NS1 RDT was also very high (100.0%; 95% CI [94.9-100.0]) and the negative agreement was very low (47.4%) with a broad 95% CI (27.3-68.3%) (Table 3). This low negative agreement correlated with the lower sensitivity of the NS1 RDT relative to RT-PCR (Table 2). Moreover, out of the 10 discordant results (NS1 RDT-negative and VIDAS^®^ NS1-positive), 7 (70.0%) were double-positive for RT-PCR and NS1 FOCUS, which both confirm an acute DENV infection [3,6,7]. This further suggests a lack of sensitivity of the NS1 RDT compared to VIDAS^®^ DENGUE NS1 Ag.

### 3.5. Comparison of VIDAS^®^ Anti-DENGUE IgM assay with competitor assays

The positive and negative agreements (95% CI) of VIDAS^®^ Anti-DENGUE IgM with the Panbio Dengue IgM Capture ELISA were 81.6% (68.6-90.0%) and 61.9% (46.8-75.0%), respectively (Table 4).

**Table 4.** Concordance of VIDAS® Anti-Dengue IgM and competitor assays.

| Reference test | Population | Positive Agreement |  | Negative Agreement |  | Overall Agreement |  |
| --- | --- | --- | --- | --- | --- | --- | --- |
|  |  | n/N | %<br>[95% CI] | n/N | %<br>[95% CI] | n/N | %<br>[95% CI] |
| **Panbio Dengue IgM Capture ELISA** | Total | 40/49 | 81.6%<br>[68.6-90.0] | 26/42 | 61.9%<br>[46.8-75.0] | 66/91 | 72.5%<br>[62.6-80.6] |
|  | Adults | 25/31 | 80.6%<br>[63.7-90.8] | 16/26 | 61.5%<br>[42.5-77.6] | 41/57 | 71.9%<br>[59.2-81.9] |
|  | Children | 15/18 | 83.3%<br>[60.8-94.2] | 10/16 | 62.5%<br>[38.6-81.5] | 25/34 | 73.5%<br>[56.9-85.4] |
| **IgM RDT<sup>a</sup>** | Total | 31/36 | 86.1%<br>[71.3-93.9] | 26/47 | 55.3%<br>[41.2-68.6] | 57/83 | 68.7%<br>[58.1-77.6] |
|  | Adults | 21/25 | 84.0%<br>[65.3-93.6] | 16/28 | 57.1%<br>[39.1-73.5] | 37/53 | 69.8%<br>[56.5-80.5] |
|  | Children | 10/11 | 90.9%<br>[62.3-98.4] | 10/19 | 52.6%<br>[31.7-72.7] | 20/30 | 66.7%<br>[48.8-80.8] |

<sup>a</sup>SD BIOLINE Dengue Duo Rapid Test (Dengue IgG/IgM); Chi-square Test between adults and children on overall agreement: VIDAS® vs. Panbio (p=0.869) and VIDAS® vs. RDT (p=0.767); Abbreviations: CI, confidence interval; RDT, rapid diagnostic test.

The VIDAS^®^ Anti-DENGUE IgM assay was in good positive agreement with the IgM RDT (86.1%; 95% CI [71.3-93.9]), despite fewer IgM RDT-positive than Panbio IgM ELISA-positive tests (n=36 vs. 49) (Table 4). By contrast, the negative agreement of the VIDAS^®^ Anti-DENGUE IgM with the IgM RDT was low (55.3%; 95% CI [41.2-68.6]) (Table 4).

### 3.6. Comparison of VIDAS^®^ Anti-DENGUE IgG assay with competitor assays

Both the positive and negative agreements of the VIDAS^®^ Anti-DENGUE IgG with the Panbio Dengue IgG Indirect ELISA were close to 80% (76.4% and 78.9%, respectively) with wide 95% CI (Table 5). As for the anti-DENV IgM assays, the VIDAS^®^ Anti-DENGUE IgG assay was in good positive agreement with the IgG RDT (89.2%; 95% CI [75.3-95.7]), despite fewer IgG RDT-positive than Panbio IgG ELISA-positive tests (n=37 vs. 72) (Table 5). Like for the anti-DENV IgM assays, the negative agreement of VIDAS^®^ Anti-DENGUE IgG with IgG RDT was low (52.2%; 95% CI [38.1-65.9]) (Table 5). Interestingly, among the 22 discordant assays negative with IgG RDT and positive with VIDAS^®^ Anti-DENGUE IgG, 19 (86.4%) were also positive in the Panbio IgG ELISA.

**Table 5.** Concordance of VIDAS® Anti-Dengue IgG and competitor assays.

| Reference test | Population | Positive Agreement |  | Negative Agreement |  | Overall Agreement |  |
| --- | --- | --- | --- | --- | --- | --- | --- |
|  |  | n/N | %<br>[95% CI] | n/N | %<br>[95% CI] | n/N | %<br>[95% CI] |
| **Panbio Dengue IgG Indirect ELISA** | Total | 55/72 | 76.4%<br>[65.4-84.7] | 15/19 | 78.9%<br>[56.7- 91.5] | 70/91 | 76.9%<br>[67.3-84.4] |
|  | Adults | 37/46 | 80.4%<br>[66.8-89.3] | 8/11 | 72.7%<br>[43.4-90.3] | 45/57 | 78.9%<br>[66.7-87.5] |
|  | Children | 18/26 | 69.2%<br>[50.0-83.5] | 7/8 | 87.5%<br>[52.9-97.8] | 25/34 | 73.5%<br>[56.9-85.4] |
| **IgG RDT<sup>a</sup>** | Total | 33/37 | 89.2%<br>[75.3-95.7] | 24/46 | 52.2%<br>[38.1-65.9] | 57/83 | 68.7%<br>[58.1-77.6] |
|  | Adults | 27/29 | 93.1%<br>[78.0-98.1] | 14/24 | 58.3%<br>[38.8-75.5] | 41/53 | 77.4%<br>[64.5-86.5] |
|  | Children | 6/8 | 75.0%<br>[40.9-92.9] | 10/22 | 45.5%<br>[26.9-65.3] | 16/30 | 53.3%<br>[36.1-69.8] |

<sup>a</sup>SD BIOLINE Dengue Duo Rapid Test (Dengue IgG/IgM); Chi-square Test between adults and children on overall agreement: VIDAS® vs. Panbio (p=0.553) and VIDAS® vs. RDT (p=0.023); Abbreviations: CI, confidence interval; RDT, rapid diagnostic test.

### 3.7. Negative agreement of the VIDAS^®^ NS1 antigen and anti-DENV IgM and IgG assays with competitor assays conducted on healthy donor samples

To verify the specificity (false-positive rate) of the three VIDAS^®^ prototypes, the negative agreement was evaluated on a cohort of negative samples. VIDAS^®^ dengue assays are intended as an aid in the diagnosis of patients with clinical symptoms consistent with dengue infection. Accordingly, the negative agreement should be evaluated on the intended population (i.e. patients with clinical symptoms consistent with dengue infection but negative for DENV RNA, DENV NS1 antigen and anti-DENV IgM/IgG antibodies, using the reference tests). Of the 250 suspected cases of DENV infection evaluated in this study (Figure 1), only 4 were defined as negative, thus preventing an accurate calculation of the false-positive rate. Therefore, negative agreement was evaluated on a population of healthy volunteers from a region non-endemic for dengue (Table 6). A total of 51 sera of adult healthy volunteers tested negative for the three competitor ELISA (NS1 FOCUS, IgM and IgG Panbio) were evaluated with the three VIDAS^®^ prototypes. The negative agreement of VIDAS^®^ DENGUE NS1 Ag and of VIDAS^®^ Anti-DENGUE IgM was 100.0% (95% CI 93.0-100.0%) and that of VIDAS^®^ Anti-DENGUE IgG was 96.1% (95% CI 86.5-99.5%) (Table 7).

**Table 6.** Healthy donors’ characteristics (N=51)

|  |  |
| --- | --- |
| Age in years, median (range) | 32.0 (18-69) |
| Gender, N (%) |  |
| Female | 34 (66.7%) |
| Male | 17 (33.3%) |
| Source of samples, N (%) |  |
| EFS, France | 51 (100.0%) |

Abbreviation: EFS, Etablissement Français du Sang

**Table 7.** Negative agreement of the VIDAS® prototypes with the competitor ELISA tested on samples of healthy donors (N=51)

| Population | VIDAS® NS1 |  | VIDAS® IgM |  | VIDAS® IgG |  |
| --- | --- | --- | --- | --- | --- | --- |
|  | n/N | %<br>[95% CI] | n/N | %<br>[95% CI] | n/N | %<br>[95% CI] |
| Adult healthy donors<sup>a</sup> | 51/51 | 100.0%<br>[93.0-100.0] | 51/51 | 100.0%<br>[93.0-100.0] | 49/51 | 96.1%<br>[86.5-99.5] |

<sup>a</sup>Tested negative for the three competitor ELISA: NS1 FOCUS, Panbio IgM and Panbio IgG. Of note, these samples were not tested for DENV RNA (RT-PCR). Abbreviation: CI, confidence interval.

## 4. Discussion

This study investigated the concordance of three VIDAS^®^ dengue prototypes detecting DENV NS1 antigen (VIDAS^®^ DENGUE NS1 Ag) and anti-DENV IgM and IgG antibodies (VIDAS^®^ Anti-DENGUE IgM and VIDAS^®^ Anti-DENGUE IgG) with competitor ELISA and RDT assays, in a population of 91 patients with acute DENV infection.

DENV RNA and dengue NS1 antigen are the two markers of choice for the confirmation of an acute DENV infection [3,6,7]. When considering RT-PCR as gold standard for an acute DENV infection, the VIDAS^®^ and FOCUS ELISA NS1 antigen assays demonstrated a high sensitivity (85.7% and 83.9%, respectively), as well as a strong positive agreement with each other (97.6%). In comparison, the NS1 RDT showed a lower sensitivity relative to RT-PCR (72.7%), as well as a low negative agreement with the VIDAS^®^ DENGUE NS1 Ag assay (47.4%). These results are in line with reports from the literature demonstrating a good concordance between DENV NS1 antigen and RT-PCR assays in acute DENV infection, as well as a better concordance of ELISA-based (considering VIDAS^®^’s principle as ELISA-based) than of RDT-based results to RT-PCR [21]. Our data thus indicate that the VIDAS^®^ and FOCUS ELISA NS1 assays are highly and comparably sensitive for the detection of acute DENV infection, and that they are more sensitive than the NS1 RDT. Previous side-by-side comparison studies actually demonstrated a high sensitivity (>95%) of the NS1 FOCUS ELISA and a comparatively lower sensitivity (<73%) of the NS1 RDT (SD BIOLINE Dengue Duo, Dengue NS1 Ag) [20,22]. The lower sensitivity of the NS1 RDT has also been confirmed in independent studies [15,19,23,33].

An inferior sensitivity of RDT vs. ELISA has also been reported for anti-DENV IgM and IgG assays [8,12,19], including with the assays used in this study (SD BIOLINE Dengue Duo IgG/IgM, Panbio Dengue IgM Capture ELISA, and Panbio Dengue IgG Indirect ELISA) [19]. This inferior sensitivity of RDT vs. ELISA might also explain the low negative agreement of the VIDAS^®^ IgM and IgG assays with the respective IgM and IgG RDT (55.3% and 52.2%, respectively). Our data thus suggest that VIDAS^®^ anti-DENV IgM/IgG assays are more sensitive than IgM/IgG RDT. On the other hand, anti-DENV IgM/IgG RDT have been shown to have a low false-positive rate [12,15,22]. The good positive agreement of the VIDAS^®^ IgM and IgG assays with the IgM/IgG RDT suggests that the VIDAS^®^ serological assays might also present a low false-positive rate, indicative of a good specificity. That the VIDAS^®^ DENGUE IgM and IgG assays but also the VIDAS^®^ NS1 Ag assay show a good specificity is supported by their high negative agreement (96.1% to 100.0%) with the reference tests on a collective of healthy donors.

As opposed to anti-DENV IgM/IgG RDT, some IgM/IgG ELISA have been described to lack in specificity, notably due to cross-reactivity with other anti-flavivirus antibodies [3,12,19,22,23]. Higher rates of false-positive results have also been reported for the competitor IgM/IgG ELISA used as reference in this study (Panbio) [12,17–19]. Differences in specificity might explain the lower level of concordance observed between the VIDAS^®^ and Panbio serological assays (72.5% and 76.9% overall agreement for IgM and IgG assays, respectively). On the other hand, one cannot exclude an impact of the nature of the antigen used in the ELISA serological tests on assay performance, as previously suggested [8,12]. The VIDAS^®^ and Panbio anti-DENV IgM and IgG tests both employ antigens specific for the four DENV serotypes, but while Panbio assays use a pool of four recombinant antigens, VIDAS^®^ assays use a recombinant tetravalent EDIIIT2 protein, which contains the DENV-specific envelope domain III (EDIII) of the four DENV serotypes [28]. As opposed to EDI and EDII, EDIII is less conserved among flaviviruses and expected to be less cross-reactive with sera from patients infected with other flaviviruses [34]. Cross-reactivity studies will help determine the false-positive rate associated with the VIDAS^®^ anti-DENV IgM and IgG assays (manuscript in preparation).

This study presents several limitations. First, the selection criteria of acute infection (RT-PCR- and/or NS1-positive) using the NS1 FOCUS assay resulted in a low number of tests negative for NS1 FOCUS (n=9), thus providing an inaccurate (broad 95% CI) evaluation of the negative agreement with the VIDAS^®^ DENGUE NS1 Ag assay (Table 3). Second, in the absence of gold standard for serological tests, agreement analyses using reference tests that might lack sensitivity and/or specificity, result in apparent discordances that are difficult to interpret, and call for future diagnostic accuracy analyses, including on negative populations of regions endemic for DENV.

## 5. Conclusions

Altogether, this study demonstrated a high sensitivity of the VIDAS^®^ DENGUE NS1 Ag assay and a good positive agreement of the three VIDAS^®^ dengue prototypes with competitor ELISA and RDT assays, indicating overall good assay performance. Like previously shown for manual ELISA, this study suggested a superior sensitivity of the three VIDAS^®^ dengue assays over RDT. Altogether, the VIDAS^®^ dengue prototypes performed very well and appear to be suitable for routine detection of dengue NS1 antigen and anti-dengue IgM/IgG antibodies. The combined use of the three VIDAS^®^ tests offers a state-of-the-art approach for the accurate diagnosis of DENV infection [3,8,10]. Finally, a clear benefit of the VIDAS^®^ platform compared to manual ELISA is the rapidity of testing, with the major advantage over a manual RDT of the automated execution and interpretation of tests, which is well suited for its implementation in regions at risk of DENV infection.

## Data Availability

The data that support the findings of this study are available on request from the corresponding author. The data are not publicly available due to privacy or ethical restrictions.

## Supplementary Materials

Not applicable.

## Acknowledgements

The authors are grateful to Ms Agnès Foussadier (bioMérieux) for her scientific contribution and to Ms Phaithong Bounmany and Thep Aksone Chindavong (Institute Pasteur du Laos) for their technical assistance. The authors thank Dr. Anne Rascle of AR Medical Writing (Regensburg, Germany) for providing medical writing support, which was funded by bioMérieux (Marcy L’Etoile, France), in accordance with Good Publication Practice (GPP3) guidelines (http://www.ismpp.org/gpp3).

## Author Contributions

Conceptualization, M.G. and S.S.; methodology, M.G. and S.S.; investigation, M.G. and S.S.; project administration, M.G. and S.S.; supervision, M.G. and S.S.; resources, M.G. and S.S.; formal analysis, L.B.; writing – review and editing, M.G., S.S. and L.B. All authors have read and agreed to the published version of the manuscript.

## Institutional Review Board Statement

The study was conducted according to the guidelines of the Declaration of Helsinki, and approved by the Institutional Review Board of the Institut Pasteur du Laos and the National Ethic Committee for Health Research of the Ministry of Health of Lao PDR (reference 049 NECHR, dated July 26, 2012).

## Informed Consent Statement

Written informed consent was obtained from all subjects involved in the study.

## Data Availability Statement

The data presented in this study are available within the article.

